# Understanding research readiness in psychological services: mixed method findings from a cross-sectional survey pilot

**DOI:** 10.1101/2025.10.28.25338875

**Authors:** Katie Saunders, Paul Campbell, Gary Lamph, Michelle Rydon-Grange, Gemma Murphy, Ben Rogers, Eleanor Bradley, Jim Grange, Ruth Lambley-Burke, Abigail Locke, Tom Kingstone

## Abstract

**Background:** Healthcare organisations that have a research culture within their practice (e.g. evidenced based and evidence informed practice) report better outcomes for their service users. There are however reported barriers to embedding research into practice. This includes time, knowledge, motivation, ability, resources, and access to organisational support. Psychological Services within the NHS regularly embed evidence-based practice as part of the care provision for their service users. However, at present little is known of the extent, capacity, and research readiness of those that practice within this sector.

**Aim:** To understand capacity, engagement and research readiness within the psychological services team at an NHS trust within the UK.

**Methods:** Mixed methods pilot study using cross sectional survey. Collecting both quantitative and qualitative data. 35 people were recruited from a pool of 89 people who attended a psychological services webinar in April 2024. Quantitative data was analysed on a descriptive level; qualitative data was analysed using thematic analysis.

**Findings and discussion:** Results indicate the value in the use of a mixed method survey to assess research readiness and capacity to those within psychological services practice. Overall response rate was good (39%) with a 100% completion rate of all questions. Both the quantitative and qualitative data revealed that participants wish to engage in research but encounter barriers such as capacity and time. Respondents were also unsure on the level or organisational support for research engagement and activity, unsure on the pathways to secure research time (e.g. funding opportunities), and findings also illustrate issues around practitioner confidence in applying research skills. Our findings show the acceptability of assessing research readiness within psychological practice and highlight several areas of need for practitioners to facilitate full research engagement. These findings will now seed a larger more ambitious assessment of research readiness.

## Introduction

NHS England identifies research in health and social care as an important function and means through which to drive service quality standards and improve outcomes for patients and service users (NHS England, 2024). This approach is exemplified through the integration of evidence-based practice (EBP) within services across the NHS (Abela, 2023); enabling practice to not only be informed by research, but to also apply research within their practice (Melchert et al., 2023; Sackett et al., 1996; Sackett, 1997). Services that adopt EBP demonstrate increased practice performance resulting in more positive outcomes for service users (Boaz et al., 2015; Ozdemir et al., 2015; Melchert et al., 2023). A key underpinning of EBP is the need to ‘support healthcare professionals to develop research skills relevant to their clinical role and to design studies in ways which ensure delivering research is a rewarding experience, rather than an additional burden’ (Department of health and social care; UK GOVT, 2021).

Whilst there is broad acceptance of the principles of EBP, health and social care clinicians and practitioners face numerous challenges with implementation (Rousseau & Gunia, 2016). Barriers typically include lack of time, knowledge, motivation, ability, resources, and access to organisational support (Alatawi et al., 2018; Barratt and Fulop, 2016; Fry and Attawet, 2018; Royal college of Physicians, 2022). Added to these generic barriers is a lack of uniform support to research engagement across health and social care sectors. A recent evaluation of the UK Government’s funding body for health and care research (National Institute for Health and Care Research, NIHR) highlights disparities linked to certain sectors who lack historical and embedded links to research in step with aligned academic institution support, meaning some professional sectors are left behind (Burkinshaw et al., 2022). This point is further expanded when taking a research culture viewpoint. For example, medical practice has with facilitators such as in-built research career progression pathways, recognised awards associated with research engagement, and a greater sense of self-belief and confidence (Dimova et al., 2018), whereas those within broader health and care roles (Allied Health, Social Care, Clinical Psychology) have limited exposure to research post qualification (Comer et al., 2022; Smith & Thew, 2017)

Psychological services, as part of NHS multi-speciality and integrated care, play a significant role in supporting people with mental health difficulties as part of community mental health approaches and NHS talking therapies (previous known as IAPT), as well as within multidisciplinary teams who support those with learning disabilities, substance misuse issues, and those that have come into contact with justice services (Tracy et al., 2019). Psychological services encompass a variety of trained professionals from therapists, councillors, and psychotherapists within integrated care systems (Gowar et al., 2024). EBP has been shown to be an important component of psychological practice (Spring, 2007), it is often used to plan and develop treatment plans and interventions for complex patients, and the implementation of EBP has demonstrated improved service user outcomes (Cook et al., 2017; Deighton et al., 2016). In recognition of EBP benefits for practice, the British Association for Counselling and Psychotherapy (BACP) now advocate its’ role within therapeutic practice (BACP, 2024). Whilst there is clear recognition and support for the adoption of EBP within Psychological Services, the current understanding of the mechanisms that underpin EBP within psychological services is limited. Current evidence shows a lack of research integration into practice, particularly for those within mental health roles such as mental health nurses (Dickens et al., 2024). At present no evidence is available to understand research capacity and engagement in psychological services within NHS settings.

Our key aim in this current study was to assess the acceptability of applying a mixed method survey to assess research readiness, engagement, and capacity within a typical NHS psychological services population in the UK. Additional aims were to: (1) understand current research confidence within psychological services; (2) describe awareness and current research practice; and (3) map and recognise the current barriers to engaging in research activities. Such findings will be an initial step to understand current “research readiness” of psychological services as a central component of EBP and highlight where further support to the engagement and use of research is required.

## Method

### Design and setting

A cross-sectional mixed method survey design was applied within this service evaluation pilot. Participants (*n = 35*) were recruited from a convenience sample (*n = 89*) of the psychological services workforce who attended a webinar that was organised to discuss staff research interests. The webinar attendees were drawn from the larger psychological services workforce (*n = 1180*) at Midlands Partnership University NHS Foundation Trust (MPFT). As well as mental health and psychological services that provide inpatient and community support, MPFT also delivers services in other clinical areas, such as, musculoskeletal, rheumatology, sexual health, psychiatry, perinatal health, amongst others. The Trust covers two counties in the Midlands region of the UK; Staffordshire (*n* = 877,900) and Shropshire (*n* = 324,700) which includes a diverse demographic population in terms of gender, age, ethnicity, and both inner-city urban and remote rural contexts. Psychological services at MPFT offer a wide range of services for people who require support for their mental health, which ranges from both inpatient to community support for a variety of presentations and disorders, the team is made up of a variety of different roles such as clinical psychologists, assistant psychologists, and recovery workers. Psychological service runs across both health and criminal justice services.

### Ethical approach

This pilot survey was undertaken by the MPFT Department for Research and Innovation in conjunction with the Psychological Services team as a “Service Evaluation”. The purpose for this approach was twofold; firstly, to indicate the acceptability of the use of a mixed method survey to assess research readiness for MPFT psychological services staff, and secondly to provide estimations of current staff research awareness and engagement. The eventual outcome is to inform on a planned future workforce survey for all psychological services staff to formulate a research needs strategy for this workforce (i.e. research training and learning needs, organisational support structure requirements, individual practitioner research pathway support) as provided by MPFT R&I. As a result of this internal service evaluation approach no formal ethical approval was required. However, the survey procedure did follow ethical principles that included informing potential participants about the survey (purpose and content), that participation was entirely voluntary, and that any data would be fully anonymised and aggregated prior to dissemination. All participants were notified that completion and submission of their survey indicated their consent to take part.

### Recruitment and procedure

A “research interest” group was established with membership from the psychological services team and individuals from the Research and Innovation team (authors of this paper represent this group) to work collaboratively to formulate the mixed method survey as a part of broader objectives to assess and facilitate research engagement for MPFT psychological services staff. Invitations to complete this pilot survey were issued to all staff members within the psychological services who had registered to a research discussion webinar (April 2024). All staff who had signed up for the webinar (*n = 89*) received information about the survey (25^th^ March 2024) and were invited to participate (via email). All potential participants received a reminder about the survey at 3 weeks after the initial invitation (15^th^ April 2024) and received a final reminder during the webinar (24^th^ April 2024).

### Measures

The measures used in this questionnaire survey were based on the MPFT Research Readiness Questionnaire (RRQ, <u>MPFT_Research_Readiness_Questionnaire_Final.pdf</u>). The MPFT RRQ was created to assess research engagement, experience, and interest within health and care sectors and was recently applied to understand research readiness and engagement within the social care sector at MPFT (Wakefield et al 2022). All items within the MPFT RRQ are based on previous literature and validated measures used to assess individual practitioner research capability, capacity and culture that can be applied within health, allied health and social care settings (Comer at el., 2022; Cordrey et al., 2023; Holden et al., 2023; Kardash, 2000; Maltese et al., 2017). Whilst the MPFT RRQ is a generic measure of research readiness for health and care, it can also be suitably adapted for specific sector-based questions. Our team therefore made minor adaptations, such as ensuring the wording about working role included reference to psychological practice, and that a Likert scale was used to assess practitioner confidence (within the research skills section) to give a more granular assessment compared with Yes/No formatting as used in the original version. The current survey used a pragmatic approach (Allemang et al., 2022) that includes both quantitative descriptive elements and qualitative open-ended questions which facilitates both an inductive and deductive interpretation of the results. See Table 1 for the domains of the survey.

**Table 1:** Domains, questions and response options for the survey.

| Table 1: Domains, questions and response options for the survey. |  |  |
| --- | --- | --- |
| Domain | Question | Response option |
| Demographic | Age, years | Categorical (Below 25, 25-29, 30-34, 35-39, 40-44, 45-49, 50-59, 60-69, Above 69) |
|  | Gender | Categorical (Female – including trans woman, Male – including trans man, non-binary, don't know, prefer not to say) |
|  | Current post | Freeform entry |
|  | Length of time in post | Categorical (0-3 years, 4-7 years, 8-15 years, above 15 years) |
|  | Part time/ full time | Categorical (Full time, Part time) |
|  | Qualifications | Categorical/ multiple choice (Certificate, diploma, Bachelor, Masters, Doctorate) Freeform (other) |
|  | Current further education | Categorical/ multiple choice (None, Certificate, diploma, Bachelor, Masters, Doctorate) Freeform (other) |
|  | Years of practice | Categorical (0-3 years, 4-7 years, 8-15 years, above 15 years) |
|  | Size of team at trust | Categorical (Sole practitioner, below 10, 11-100, 101-1000, above 1000) |
| Existing research engagement. | Involvement in research in the past 3 years | Categorical (Yes, No) |
|  | brief description of research activity or training. | Optional (if yes to previous question) Freeform entry. |
| Importance and relevance of research. | Research as a part of professional development | Categorical (Yes, No) |
|  | Research as a part of professional development | Open ended, following on from previous question. |
|  | Relevance of research to current practice. | Categorical (not at all, slightly, moderately, very, extremely). |
|  | Relevance of research to current practice. | Open ended, following on from previous question. |
|  | Value of research to psychological services. | Categorical (No value, minimal value, quite valuable, extremely valuable). |
|  | Value of research to building relationships with other healthcare professionals. | Categorical (No value, minimal value, quite valuable, extremely valuable). |
|  | Value of research to maintaining interest in psychological services. | Categorical (No value, minimal value, quite valuable, extremely valuable). |
| Knowledge/ Interest of/ In research. | Up to date with existing literature. | Categorical (not at all, slightly, moderately, very, extremely). |
|  | Interest in conducting own research. | Categorical (not at all, slightly, moderately, very, extremely). |
|  | Current research interests. | Open ended, freeform. |
| Organisational research support. | Does your organisation have... |  |
|  | Adequate resources to support staff research training. | Categorical (Yes, no, Partly, I don't know) |
|  | Senior managers that support research. | Categorical (Yes, no, Partly, I don't know) |
|  | Ability to ensure that career pathways are available in research. | Categorical (Yes, no, Partly, I don't know) |
|  | Ability to ensure organisation practice is guided by evidence. | Categorical (Yes, no, Partly, I don't know) |
|  | Software programmes for analysing research data. | Categorical (Yes, no, Partly, I don't know) |
|  | Support with applications for research scholarships/ degrees. | Categorical (Yes, no, Partly, I don't know) |
| Confidence in research skills | A. Finding relevant literature.<br>B. Critically appraising research literature.<br>C. Designing a research study.<br>D. Writing a research protocol.<br>E. Collecting data e.g., surveys, interviews.<br>F. Analysing qualitative research data.<br>G. Analysing quantitative research data.<br>H. Using computer data management and analysis programmes (e.g., SPSS, STATA, NVIVO)<br>I. Writing a research report.<br>J. Writing for publication in peer reviewed journals.<br>K. Submitting an ethics application.<br>L. Applying for research funding.<br>M. Writing a research abstract, submitted for a psychological service-related conference.<br>N. Overall ability to design or conduct research. | Categorical (1. No confidence, 2. Little confidence, 3. Moderately confident, 4. Very confident, 5. Extremely confident) |
| Participants questions about research. | What question would they ask as a psychological services researcher. | Open ended, freeform |
|  | Participants relationship to research in their practice. | Open ended, freeform |
|  | Key barrier or barrier to involvement and engagement in research. | Open ended, freeform |

### Analysis

Microsoft Forms was used to collect the survey data from each participant. The raw data was subsequently downloaded into Microsoft Excel file. The quantitative data and qualitative data were then separated, and the quantitative data imported into SPSS (version 29) for descriptive analysis. Some questions within the survey offered an option for ‘*Other’* with an open response box, for example ‘*Does your organisation have the ability to ensure staff career pathways are available in research*?’ with answers being *‘yes’, ‘no’, ‘partly’, ‘I don’t know’, ‘other – free text box’* to support with analysis the text reported in the free text box were labelled as ‘other’. Open response boxes were also offered for the ‘Organisational research support’ section of the survey. Responses that were detailed in these boxes related to people not knowing what services their organisation offered so these were grouped under the ‘I don’t know’ Likert response option. Descriptive quantitative data analysis was applied to categorical and Likert scale questions to produce percentage proportions. No inferential statistical tests were applied due to the nature of this pilot survey and low sample size.

The qualitative data from open ended survey questions was analysed using a thematic approach (Braun & Clarke, 2006). This approach supported the identification of patterns in the data and common themes between responses. Coding was conducted by KS and remained at a descriptive/semantic level due the nature of the collected data. This summary approach was taken as the use of singular open-ended questions within a survey format offers no option for further exploration and probing as would be found within more traditional “interview” designs (LaDonna et al., 2018). Themes were agreed through discussion with experienced qualitative researchers (TK, GL, PC).

## Results

### Quantitative descriptive analysis

In total 35 participants completed the survey from an eligible population of 89 attendees at the webinar; this represents a 39% response rate for this pilot. There was no “missing” or incomplete responses within the questionnaire, and no comments about the questionnaire raised by respondents, indicating acceptability and suitability of the questions within this population. This information has been informative to our teams plans to run the survey across the entire MPFT psychological services workforce (*n = 1180*) at a future date. Table 2 presents the demographic information of the respondents.

**Table 2.**
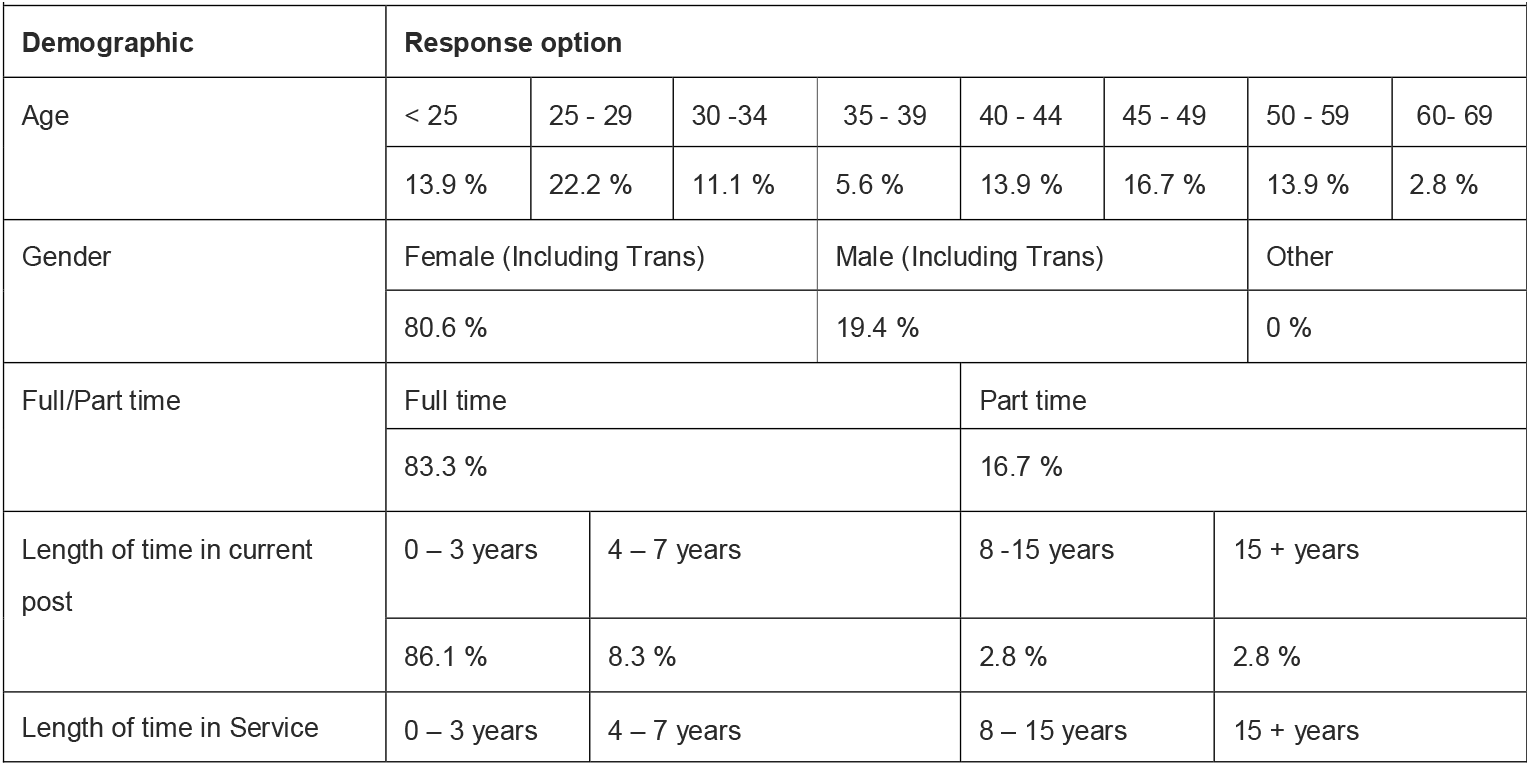
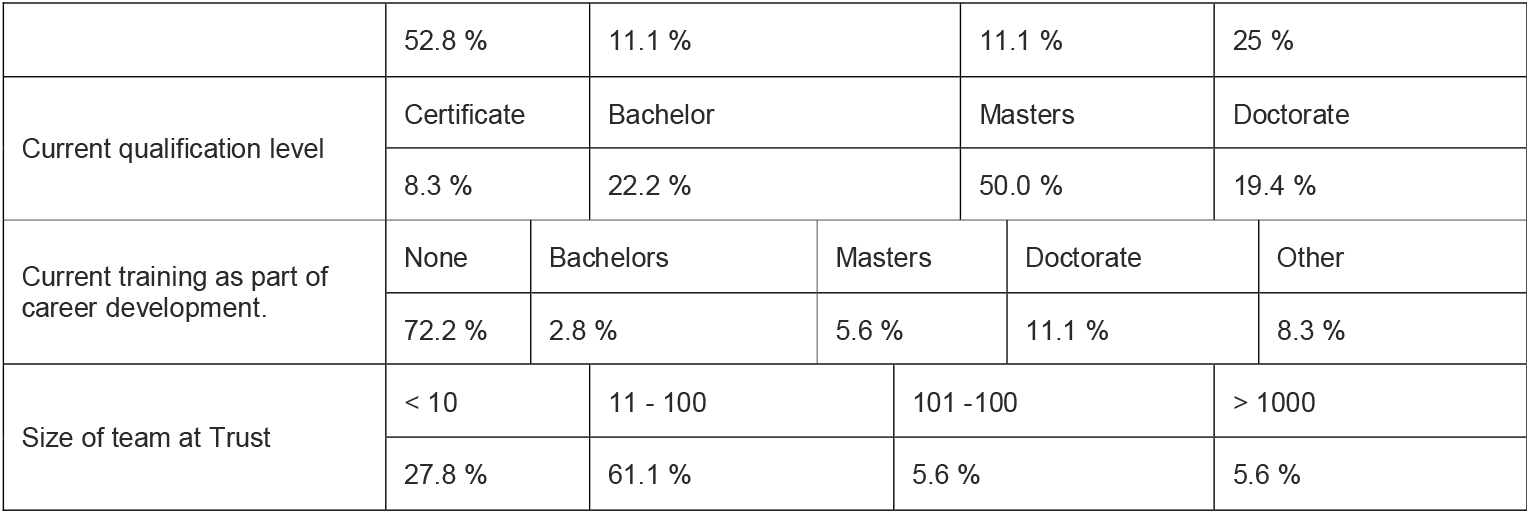
Descriptive data and demographics of the respondents.

| Demographic | Response option |  |  |  |  |  |  |  |
| --- | --- | --- | --- | --- | --- | --- | --- | --- |
| Age | < 25 | 25 - 29 | 30 -34 | 35 - 39 | 40 - 44 | 45 - 49 | 50 - 59 | 60- 69 |
|  | 13.9 % | 22.2 % | 11.1 % | 5.6 % | 13.9 % | 16.7 % | 13.9 % | 2.8 % |
| Gender | Female (Including Trans) |  |  | Male (Including Trans) |  |  | Other |  |
|  | 80.6 % |  |  | 19.4 % |  |  | 0 % |  |
| Full/Part time | Full time |  |  |  | Part time |  |  |  |
|  | 83.3 % |  |  |  | 16.7 % |  |  |  |
| Length of time in current post | 0 – 3 years | 4 – 7 years |  |  | 8 -15 years |  | 15 + years |  |
|  | 86.1 % | 8.3 % |  |  | 2.8 % |  | 2.8 % |  |
| Length of time in Service | 0 – 3 years | 4 – 7 years |  |  | 8 – 15 years |  | 15 + years |  |
|  | 52.8 % |  | 11.1 % |  | 11.1 % |  | 25 % |  |
| Current qualification level | Certificate |  | Bachelor |  | Masters |  | Doctorate |  |
|  | 8.3 % |  | 22.2 % |  | 50.0 % |  | 19.4 % |  |
| Current training as part of career development. | None | Bachelors |  | Masters |  | Doctorate |  | Other |
|  | 72.2 % | 2.8 % |  | 5.6 % |  | 11.1 % |  | 8.3 % |
| Size of team at Trust | < 10 |  | 11 - 100 |  | 101 -100 |  | > 1000 |  |
|  | 27.8 % |  | 61.1 % |  | 5.6 % |  | 5.6 % |  |

In brief, the sample was predominately female (80.6 percent), with the largest representation within the age groups 25 - 29 years (22.2%). Most staff described their role as full time (83.3%), 86.1% have been in their current role for zero to three□ years. Most participants reported that they had a master’s degree (50.0%), with most respondents not currently undertaking any further research training or qualifications (72.2%).

Table 3 displays research importance, knowledge, and skills of the participants. All respondents (100%) indicated that research should be part of their professional development with more than half (63.9%) indicating involvement in research activity or training within the last three years (63.9%). The majority of participants felt that research was extremely relevant to their current field of practice (66.7%) and also valued research for the future of psychological services (88.9%). Most participants felt research was ‘extremely valuable’ to building relationships with other healthcare professionals (69.4%). There were mixed reports in terms of how up to date participants felt with research-based theory and practice, with 42.7% saying they were moderately up to date and 33.3% saying they were very up to date. The overwhelming majority (91.7%) indicated they would be “very” or “extremely” interested in conducting their own research as part of their career development.

**Table 3.**
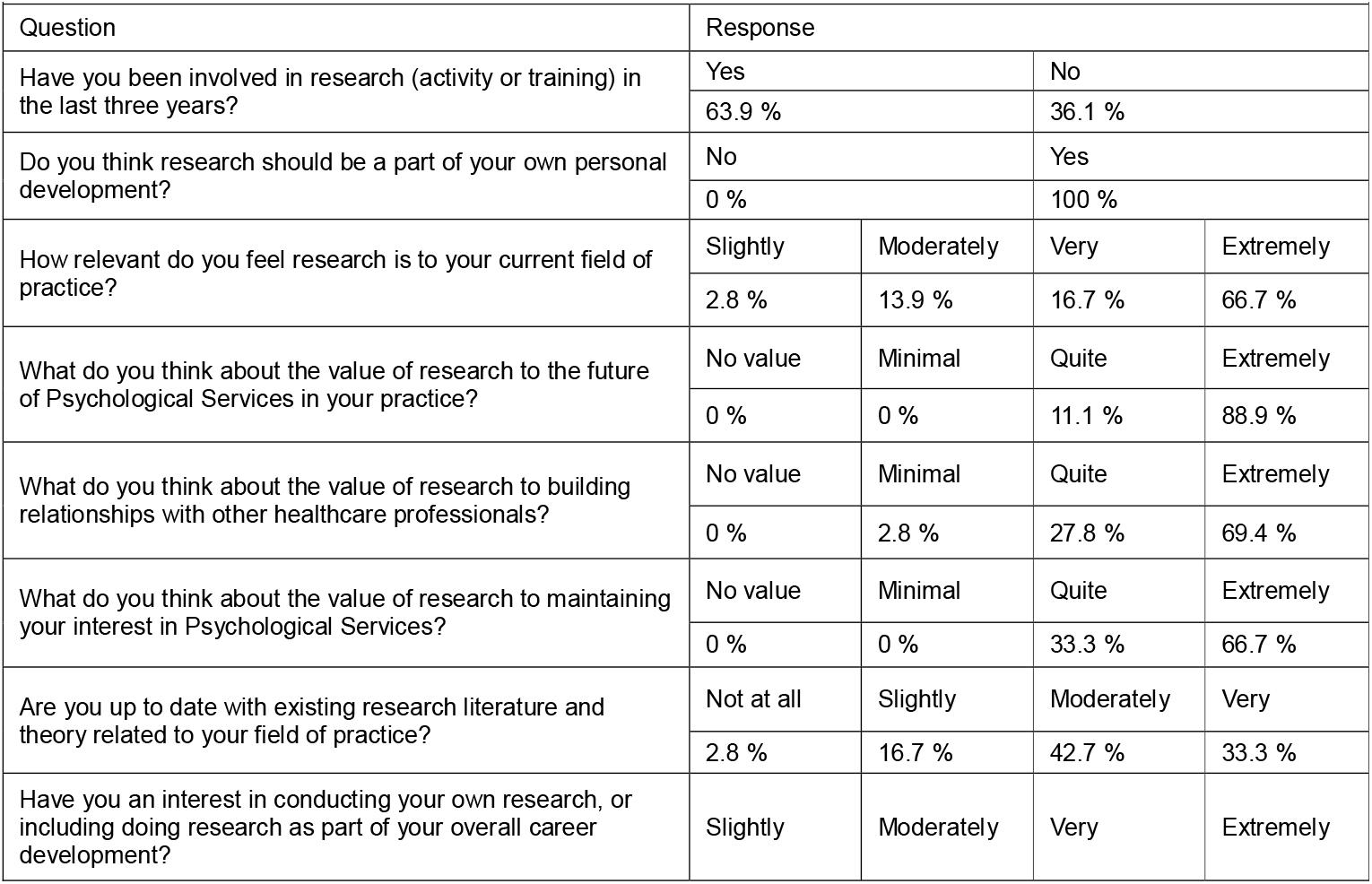
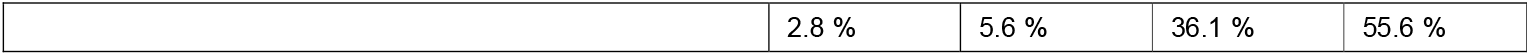
Reported importance, knowledge, and interest in research.

Respondents reported opinions about the research support in their current organisation (see table 4). There was a split across participants when it came to whether their organisation has adequate resources to support research training (yes 27.8%; partly 33.3%, don’t know 30.6%), with more than half indicating they did not know if their organisation provides advice about career pathways in research (58.3%). More than half the participants stated that yes, their organisation ensures practice is guided by evidence (55.6%), though two thirds (66.7%) were unsure on the provision of software programmes, with half (50%) of respondents being unaware of any support for research scholarship.

**Table 4.** Reported organisational research support.

| Table 4 – Reported organisational research support. |  |  |  |  |
| --- | --- | --- | --- | --- |
| Question – does your organisation have/ provide... | Yes | No | Partly | Don't know |
| Adequate resources to support staff research training | 27.8 % | 8.3 % | 33.3 % | 30.6 % |
| Ability to ensure staff career pathways are available in research. | 22.2 % | 8.3 % | 11.1 % | 58.3 % |
| Ability to ensure organisation practice is guided by evidence | 55.6 % | 2.8 % | 25 % | 16.7 % |
| Software programs for analysing research data? | 8.3 % | 13.9 % | 11.1 % | 66.7 % |
| Support with applications for research scholarships/degrees? | 16.7 % | 13.9 % | 19.4 % | 50 % |

Table 5 details the participants confidence in research skills. When it came to overall confidence in research skills, most participants (52.8%) reported that they were quite confident or had full confidence in their abilities. However notable markers of no, not very or moderate confidence were, applying for research funding (88.8%), using data analysis software (69.4%), submitting for ethics approval (66.7%), designing a research protocol (66.6%), and writing for a publication (61.1%).

**Table 5.**
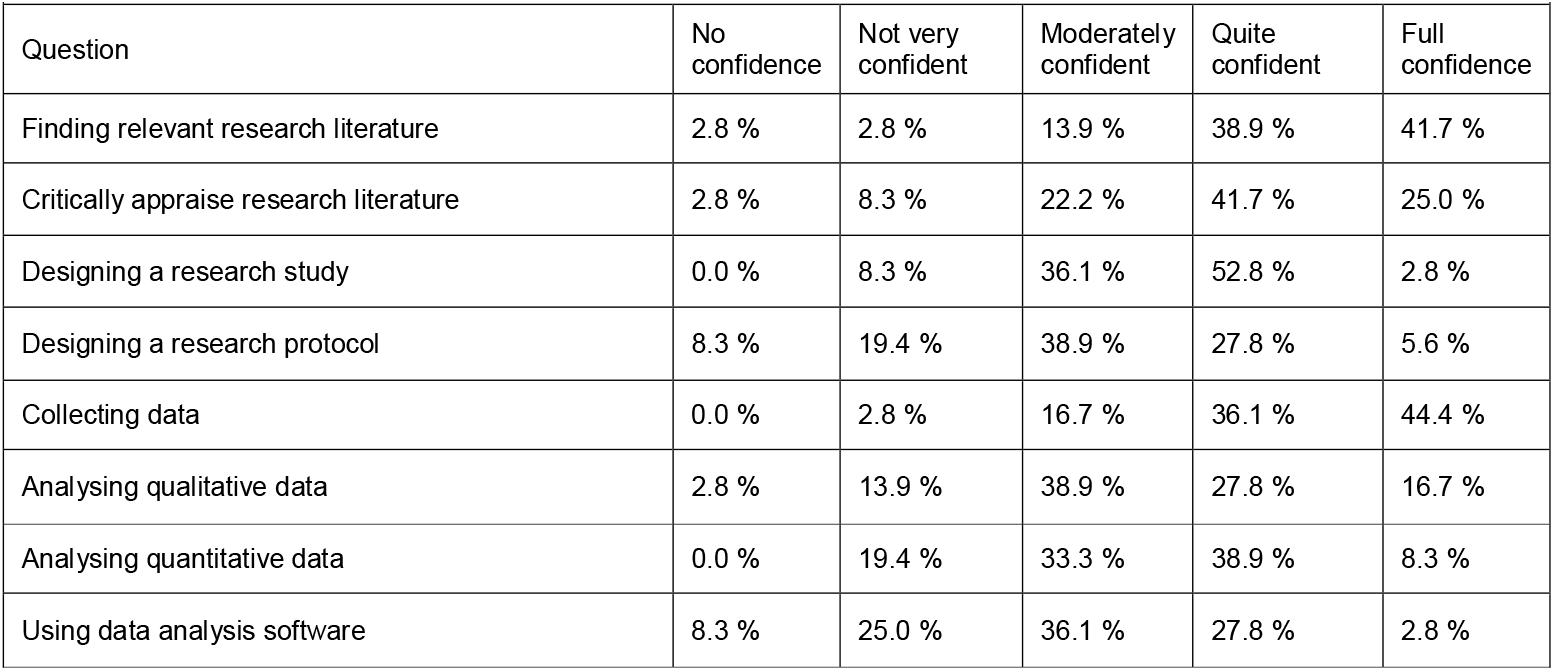
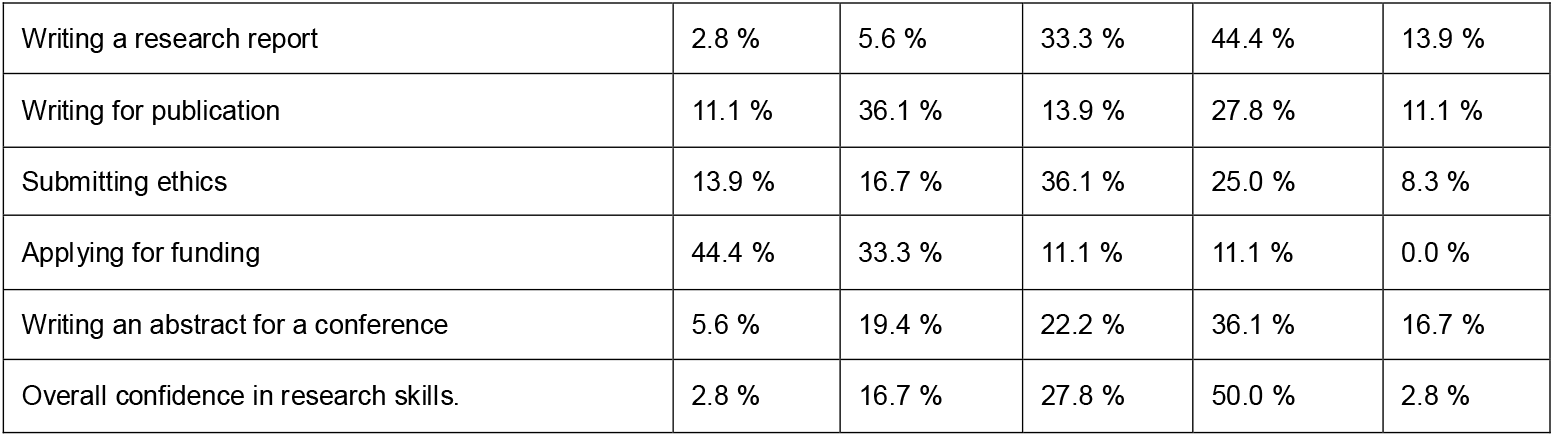
Reported confidence in research skills.

| Table 5 – Reported confidence in research skills |  |  |  |  |  |
| --- | --- | --- | --- | --- | --- |
| Question | No confidence | Not very confident | Moderately confident | Quite confident | Full confidence |
| Finding relevant research literature | 2.8 % | 2.8 % | 13.9 % | 38.9 % | 41.7 % |
| Critically appraise research literature | 2.8 % | 8.3 % | 22.2 % | 41.7 % | 25.0 % |
| Designing a research study | 0.0 % | 8.3 % | 36.1 % | 52.8 % | 2.8 % |
| Designing a research protocol | 8.3 % | 19.4 % | 38.9 % | 27.8 % | 5.6 % |
| Collecting data | 0.0 % | 2.8 % | 16.7 % | 36.1 % | 44.4 % |
| Analysing qualitative data | 2.8 % | 13.9 % | 38.9 % | 27.8 % | 16.7 % |
| Analysing quantitative data | 0.0 % | 19.4 % | 33.3 % | 38.9 % | 8.3 % |
| Using data analysis software | 8.3 % | 25.0 % | 36.1 % | 27.8 % | 2.8 % |
| Writing a research report | 2.8 % | 5.6 % | 33.3 % | 44.4 % | 13.9 % |
| Writing for publication | 11.1 % | 36.1 % | 13.9 % | 27.8 % | 11.1 % |
| Submitting ethics | 13.9 % | 16.7 % | 36.1 % | 25.0 % | 8.3 % |
| Applying for funding | 44.4 % | 33.3 % | 11.1 % | 11.1 % | 0.0 % |
| Writing an abstract for a conference | 5.6 % | 19.4 % | 22.2 % | 36.1 % | 16.7 % |
| Overall confidence in research skills. | 2.8 % | 16.7 % | 27.8 % | 50.0 % | 2.8 % |

### Qualitative analysis

Qualitative analysis revealed four themes (1) Recognising the broad relevance of research to psychological services, (2) Lack of time, capacity and support to engage in research, (3) Proactively engaging in research, and (4) Identifying research support needs, interests and ambitions. Findings in relation to theme 1, revealed that participants believed that research was vital to their role, and improved service user outcomes. This was indicative, due to links with evidence-based practice; and they often kept up to date with current research in their area of interest. Theme 2 highlights the current barriers that psychological services face in relation to research engagement. Participants discussed that time and capacity were key barriers as well as resources, skills, and organisational support. Extracts related to theme 3 highlighted how participants are either engaged or are attempting to engage in their own research, but this action is intertwined by thoughts that research can be overwhelming leading many to feel unsure. To overcome this, participants stated that they often initiate conversations around research in their practice and recognise the need to seek training. Finally, in part theme 4 encompasses answers from the question “What question would they ask to a psychological services researcher?”. Participants wanted to know how researchers strike a balance with their clinical work, how to begin a project, as well as questions relating to methodological standpoints. Participants also highlighted their research interests as part of theme 4, mainly encompassing interests, typically align with their roles, or for service improvement. Table 6 details the qualitative themes, concepts, and example texts.

**Table 6.**
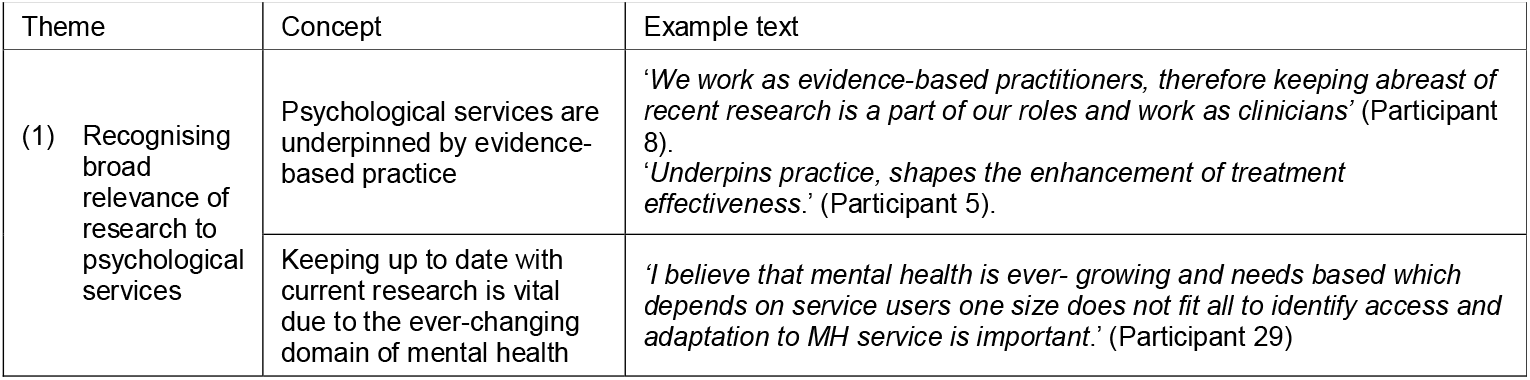
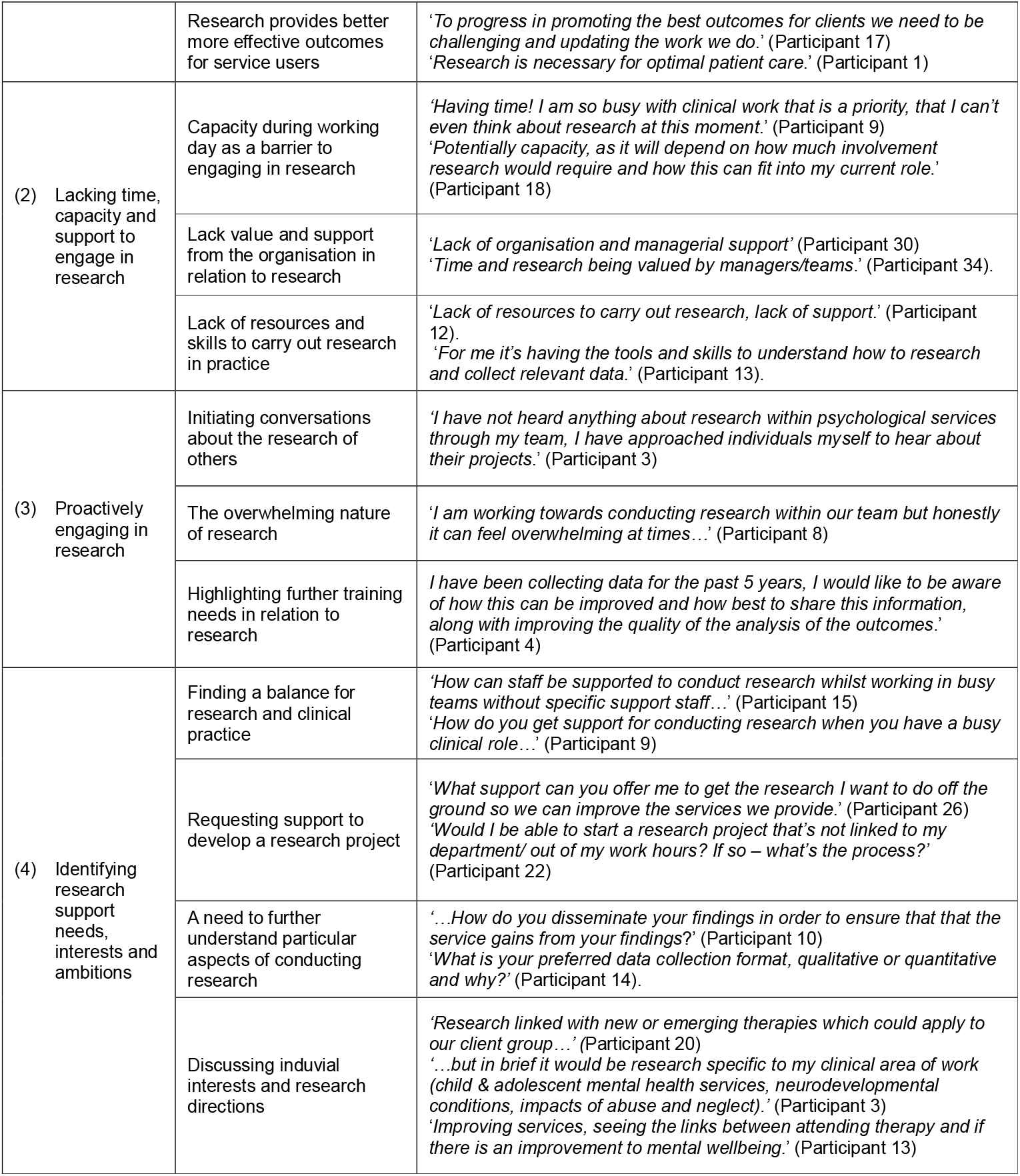
Qualitative data: Including themes, concepts and example text extracts.

## Discussion

### Overall findings and comparison with previous literature

This pilot service evaluation demonstrated the acceptability and feasibility of a survey method to assess research readiness within a psychological services workforce. There was an overall good response rate to the survey, and all respondents completed all questions. The results from the survey also are informative by giving detail to the research engagement and readiness of MPFT psychological services staff. Participants indicated that research is vital to, and promotes better outcomes, for evidence-based practice. Such findings accord with existing research that reports similar outcomes relating to effectiveness of EBP across varied health and care sectors (Boaz et al., 2015; Jonker et al., 2020; NHS England, 2024; Ozdemir et al., 2015), and more specifically research being vital to current psychological practice (BACP, 2024). Overall findings can also relate to motivations that promote self-development in research skills and embedding research into current practice (Dimova et al., 2018).

Participants provided insight into their current confidence levels in relation to research specific activities, with varying levels of confidence. When focusing on individual components there are areas that show low levels of confidence such as knowledge on how to apply for research funding, use of data analysis software, submitting a study for ethical approval, designing a research protocol and writing for publication. All of these areas could form targets for future training and learning provisions. Subject to confirmation of these results within the planned full workforce or national survey).

Regardless of individual skill confidence and a willingness to engage with research, a key finding is the reported barriers and limitations to being involved and undertaking research. These outcomes support previous research which suggests that time, resources, and knowledge are barriers to research engagement. Time was a key highlighted barrier in the current study, detailing how some may have to extend their research activities outside of their contracted hours as has been illustrated in other research (Barratt & Fulop, 2016; Fry & Attawet, 2018; Mustafa et al., 2018; Royal college of Physicians, 2022). Organisational support and resources were also inferred as a barrier in the current study; aligning with similar findings noted in similar evaluation studies at MPFT (Tajuria et al., 2024, Wakefield et al., 2022).

### Strengths and limitations

There are noted strengths associated with the current study. At present we are not aware of any other research that has explored and mapped research engagement within an NHS based psychological service setting in the UK. This pilot survey also included an acceptable response rate (39%) which is relatively higher to other similar surveys of research engagement (Friesen & Comino, 2017; Matus et al., 2019). Furthermore the 100% completion rate for all questions (i.e. no missing data) indicates the acceptability and relevance of the questions for this population and the suitability for a planned larger scale survey. Whilst a favourable response rate has been reported within the pilot, we cannot rule out response bias as the webinar attendees (from which we recruited from) were attending a webinar about research and the total attendee number (n = 89) only represents 3% of the psychological service workforce at MPFT. In addition, due to this potential bias (i.e. those taking part had a greater interest in research) the results reported here may well be under and over estimations. A further strength of this study is the use of a mixed method questionnaire as this allows for a breadth (scale and scope) and depth (understanding). However, there are limitations associated with the use of qualitative open ended response items within questionnaires. Whilst useful they are still restrictive as there is no opportunity to probe and expand on participants responses, therefore further elaboration could not be explored, which can be attained via methods such as interviews (LaDonna et al., 2018). Another general limitation is the low sample size as this restricts the ability to undertake inferential analysis. For example, a larger sample would allow testing of the characteristics associated with high or low research engagement which could identify specific areas for intervention development. Related to this point is the restrictiveness of the cross-sectional design as this does not give an understanding of potential trend and change points, ideally a longitudinal approach would offer greater information in this regard. Overall, the current study did provide further insight into engagement and research readiness within the psychological services as well as highlighted barriers and areas for service improvement.

### Implications

The findings from the current study demonstrate the feasibility and acceptability in the use of a mixed method survey to assess psychological services research readiness and engagement. This current study will be instrumental in the application of a larger scale survey which will enable for much more in-depth analysis to understand the barriers and facilitators of research engagement. In turn such information will inform our trust (MPFT) on where further support, training, learning, and organisational structural change is required to ensure all practitioners have confidence and opportunity to apply evidenced based practice to its’ fullest extent. This is clearly wanted by practice as evidenced by the high percentage of participants feeling that research it vital to their practice, and every respondent believing that research should be part of their career development. Certainly, our results here show that a lack of interest in research is not the key issue, more so a lack of understanding of the opportunities to upskill and the pathways available to undertake research career development. Such issues can be addressed by informing practice of these opportunities, for example MPFT offers a comprehensive research training and development programme called STARS (<u>STARS - The Research and Education Programme :: Midlands Partnership University NHS Foundation Trust</u>) and the R&I Department can assist and guide individual practitioners who wish to explore and undertake research career development or research activity (via funding application processes). However, an important finding from this study is the organisational barriers that research training, advice, and guidance cannot alone overcome. This central issue is practitioner time to engage with research. It has been demonstrated previously that research engagement post registration can be difficult due to clinical constraints (Comer et al., 2022; Smith & Thew, 2017) and there is a need to ensure from an organisational standpoint that dedicated time for research is embedded within normal practice. Such commitment takes vision and investment, but as evidence shows health and care organisations who adopt a research ethos and culture demonstrate improvements for both practice and patients/service user outcomes (Djulbegovic & Guyatt, 2017; Lugtenburg et al., 2009; Spring, 2007). MPFT psychological services fully endorse and apply evidenced based practice and have now taken initial steps to foster greater practitioner research engagement as an important component to this approach.

## Data Availability

All data produced in the present study are available upon reasonable request to the authors

## Acknowledgements

Psychological services staff members at Midlands Partnership University NHS Foundation Trust for taking part in the survey.

## Funding

There was no funding associated with this project.

## Notes

### Competing Interest Statement

The authors have declared no competing interest.

### Funding Statement

This study did not receive any funding

### Author Declarations

Ethics committee/IRB of Midlands Partnership University NHS Foundation Trust waived ethical approval for this work

### Summary of Updates

Author Abigail Locke was unintendedly missed off the authors list so we have added them onto the manuscript and the list of authors.

